# COVID-19: A model correlating BCG vaccination to protection from mortality implicates trained immunity

**DOI:** 10.1101/2020.04.10.20060905

**Authors:** Cameron M. Green, Stephanie Fanucchi, Jorge Dominguez-Andres, Ezio T. Fok, Simone J.C.F.M. Moorlag, Yutaka Negishi, Leo A. B. Joosten, Mihai G. Netea, Musa M. Mhlanga

## Abstract

- We use a data quality model to demonstrate that BCG vaccination is correlated with protection from death from COVID19
- From a mechanistic perspective, BCG is well described to elicit its protective non-specific effects through the process of trained immunity.
- Therapeutically enhancing trained immunity may therefore be an important mechanism in protection from the lethal effects of COVID19

## Introduction

The emergence of the COVID-19 pandemic has spotlighted the susceptibilities of different populations to its most lethal effects. Based on our current knowledge, no human being on Earth has previously been exposed to SARS-Cov2, the causal virus of COVID-19 disease. As a consequence, the role of adaptive immunity becomes more subordinate to innate immune responses. Thus, early protection from the most lethal effects of COVID-19 are more a function of the non-specific effects of the immune system rather than the acquisition of long term adaptive immunity.

Several lines of evidence have suggested that the Bacille Calmette-Guérin (BCG) vaccine exerts such non-specific effects, especially in offering protection from non-mycobacterial respiratory tract infections.^1,2^ This motivated us to more deeply investigate whether BCG provides protection from disease or death in individuals infected with SARS-Cov2. Indeed, what is unanswered is *how* BCG would confer such non-specific protection from death from COVID-19. Furthermore, if BCG does confer protection, how durable is this effect and can it be quantified? However, prior to being able to attribute and further investigate a protective role to BCG vaccination, a *bona fide* correlation between vaccination and protection from death from COVID-19 must first be established.

Seen differently, the global COVID-19 pandemic is essentially a large-scale, uncontrolled experiment on the non-specific effects of a widely used vaccine. Thus, we sought to interrogate the ecology in which the COVID-19 pandemic is occurring. Importantly, we modeled the available data from the point of view of data quality and data science and less than that of epidemiology, using what would be termed an “ecological” approach. Epidemiology requires a longer duration of a pandemic to collect useful and robust data amenable to modeling with epidemiological tools.^3^ Clearly the complete tool kit for the epidemiological approach is unavailable at this early stage.

A significant challenge early in the COVID-19 pandemic has been identifying available “clean” data.^4^ Multiple lines of evidence underline the poor quality of the majority of available COVID-19 data, and in particular the poor quality of comparable “case data” which is highly influenced by a country’s testing at this stage.^5^ This significantly influences the ability to accurately calculate case fatality rate (CFR)^6^ and as a consequence, CFR estimations are unreliable for the purposes of establishing *bona fide* correlations. Importantly, exponential growth, particularly in the early stages of this rapidly changing event, compounds these errors due to poor quality data over time.

The absence of usable tools and a paucity of trustworthy data for COVID-19 are in stark contrast to those available for SARS and MERS, for example. With *hindsight* those epidemics can be well studied and dissected using the state-of-the-art tools. However, in a fast moving and exponentially expanding disease such as COVID-19, data quality enabled data science may contribute beneficially *early-on* with *foresight* to public health policy and scientific approaches.

In the present study we use mortality data from the COVID-19 pandemic as the most robust data available at this point in time. In correcting for the temporal nature of the pandemic, with deaths initiating at different times in different countries, we used continuous rolling snapshots of deaths per million over time. This analysis was conducted for the top 100 countries ranked by mortality per capita, and normalized in time at 0.1 deaths per million for each country. Using this as a point of departure to construct bona fide correlations, we proceeded to develop a high confidence model for a group of the approximately two dozen countries with the largest range of higher quality mortality data. This analysis clearly demonstrated *whether* BCG vaccination is correlated with protection from death from COVID-19. We then linked this correlation to new and existing studies and data suggesting causal protective mechanisms that have been previously and rigorously demonstrated to be mediated by BCG vaccination. These mechanisms converge on “trained immunity,” suggesting that therapeutically enhancing trained immunity may be an important mechanism in individuals demonstrating protection from the lethal effects of COVID-19.

## RESULTS

### Establishing a robust correlation between BCG vaccination and protection from death from COVID-19

Science currently suffers from an undersupply of quality data and an oversupply of models at this early stage in the pandemic. To “see around corners”, requires multiple *trustworthy* observational points that can be used to analyze and develop robust correlations. These data should be sufficient in quantity and quality to plot a *bona fide* function. To understand the “ecology” of BCG vaccination in general, multiple temporal data points must be taken in places where BCG vaccination is routine and then compared to geographies where this does not occur.

To do so we surmised that “mortality data” was the most robust data available today. We arrived at this conclusion through several routes. Firstly, and perhaps most significantly, is the general inconsistency in how the case data is collected and reported across time within nations themselves as priorities and resources change from week to week. For example, before sufficient test resources were available in many European countries, the early priority was to test the very ill only, followed by medical workers and then suspected cases. Contrastingly, in Asia the testing early on was focused on containment and thus testing was prioritized for contacts of confirmed cases, before rapidly being moved on to the wider population. Secondly, the sheer number of tests completed to date varies from country to country. Countries that are motivated and have the means to efficiently conduct high numbers of tests per day influence the case numbers heavily in one direction while several large countries still have low testing volumes, making comparisons between countries highly uneven. Thirdly, there is a large amount of inconsistent reporting of the confirmed case data itself, for example asymptomatic individuals were not reported in some countries and this further weakens the reliability of the number of positive cases for comparison. Fourthly, in the early stages of a pandemic testing is neither free of charge, nor widely available except (with important exceptions) in the highest income countries skewing comparisons of testing data across countries in time while resources lag. Lastly, and perhaps a hard to resolve problem with testing data, is that it remains highly political as a metric, influencing its transparent reporting in many places.

Consequently, case-fatality rates (CFR), the ratio of deaths to confirmed cases, remain unreliable for comparisons as the denominators are highly dependent on test penetration per country.

In summary, test and case data is highly useful in other contexts. However, for the aforementioned reasons it is fundamentally unreliable in developing a robust correlative relationship between BCG vaccination countries and protection from death from COVID19.

“Mortality data” also has its flaws. Countries have had varying definitions for COVID-19 mortality. In some cases, a death with obvious symptoms was considered in scope, whilst for others a positive test had to be produced. Some countries in Europe did not initially include any deaths outside hospitals but these were subsequently incorporated. Countries have varying healthcare death certification protocols and timings, so some data compresses in days but is normally resolved timeously for total deaths. Also, there is concern that some deaths go unreported as health systems become saturated. However, in countries in the midst of the pandemic, abnormal spikes in numbers of deaths are noticeable and usually well reported in the press or in all-cause mortality checks where post-hoc attribution to COVID-19 can be made.

Taking the above into account, to construct our model we followed mortality data, reversing in time to early in the beginning stages of the COVID-19 outbreak in each country. Starting at the point at which each country reached 0.1 deaths per million inhabitants (**Supplemental Table 1**), we plotted cumulative deaths per million against time in days (**Figure 1 & Supplemental Table 2**). The long-term asymptote on this function reflected deaths per million population for the current wave. Examining this data for countries which started their epidemic earlier, for example China, Japan and South Korea, resulted in sufficient numbers of data points plotted to present day to be able to derive an approximate function from which we could deduce longer term COVID-19 severity per country for comparison. However, for many countries there were simply too few data points in time after the data point when 0.1 deaths per million inhabitants occurs to derive a suitable progression curve. For example, for Brazil there were only 14 such points of data because we are early in their outbreak, and obtaining a longer-term function on so few points of data is not generally predictive, yielding low confidence (**Table 1**). However, by choosing an initiation point of the crisis amenable to the generation of an approximate progression curve for as many countries as possible, and basing our plots and temporal snapshots only on the higher quality deaths per capita, we were able to distill interesting empirical results (**Figure 1**).

**Figure 1.**
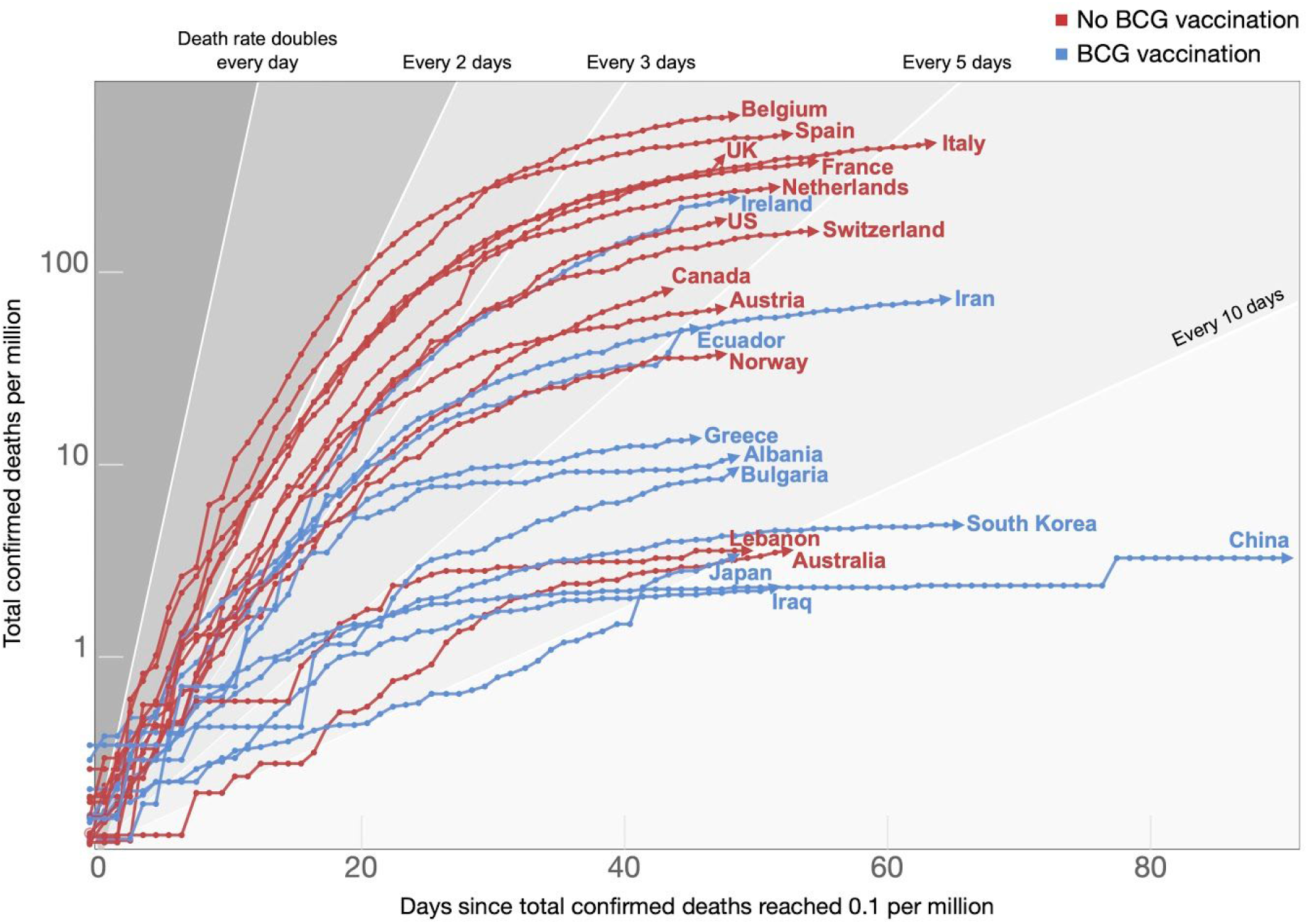
In general, countries with an ongoing BCG vaccination program exhibit reduced COVID-19-associated mortality rates. Data represents days since the total confirmed deaths of COVID-19 per million people reached 0.1. The countries represented here were selected as they have available data for > 35 days after 0.1 deaths per million. The variable time span is from 31 December 2019 to 31 April 2020. Potential explanations for outliers (such as Ireland, Australia and Lebanon) are explained in supplemental data. Source data^4^.

**Table 1.**
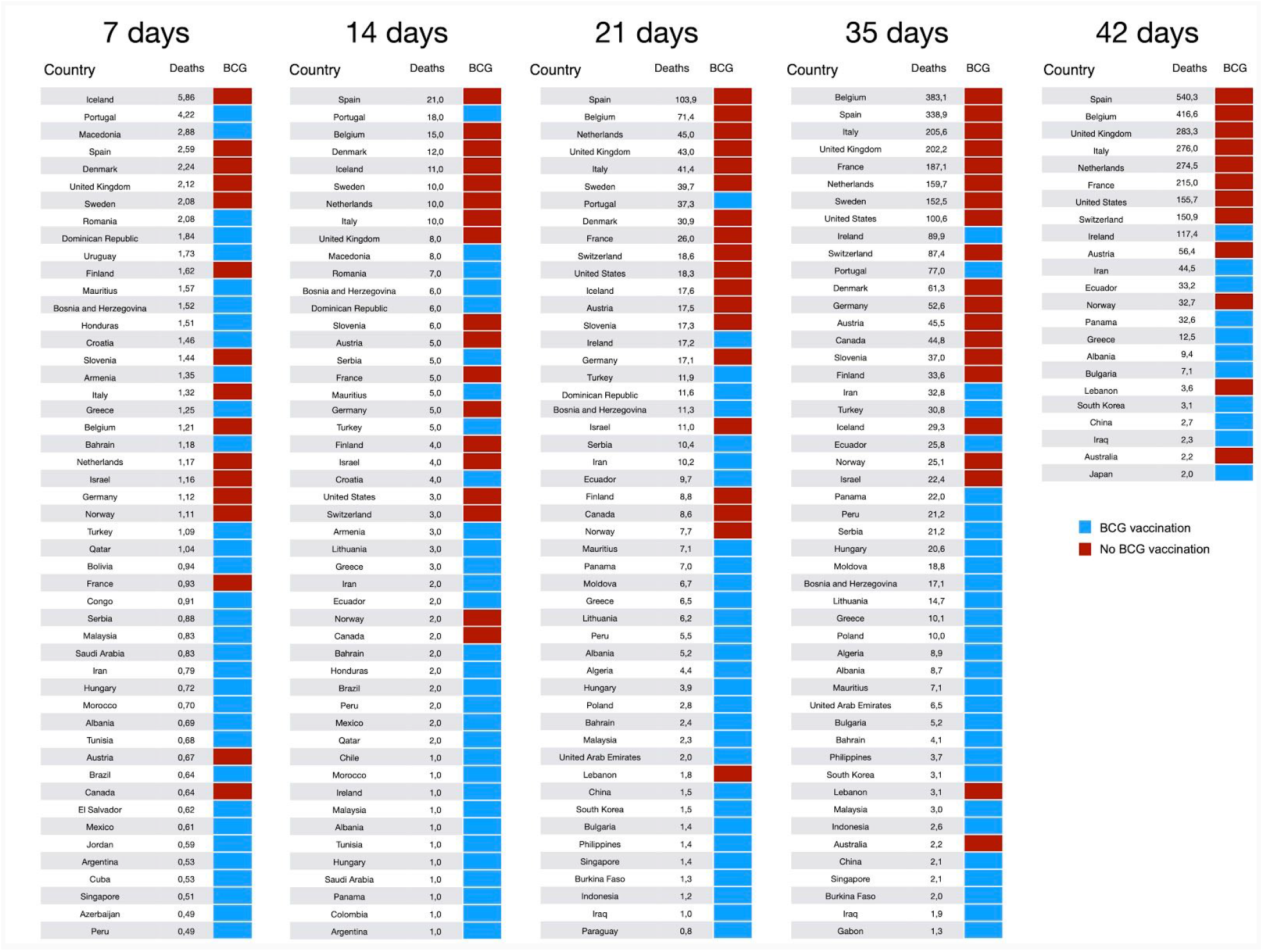
Mortality rankings per country at 7, 14, 21, 35 and 42 days since total confirmed deaths reached 0.1 per million. Increasing confidence in correlations is observed over time, with a greater number of countries without current BCG vaccine programs entering the higher ranks over time.

Noise in the per capita data can occur if countries below a million inhabitants are included. Therefore, for the purposes of this analysis, small nation states, that is nations with populations below a million people, are excluded. As previously mentioned, when this analysis is conducted, only for a small number of countries can multiple snapshots in time be obtained for up to 42 data points (**Figure 1 & Table 1** last column). This number of data points affords us a comparable semi-log graph of deaths per million data for a reasonable number of countries. Also, after preparing the table data in this way and studying the relative placement of each country per time snapshot, the model partitions well, BCG vaccination countries from those where no BCG vaccination is occurring or has been halted (**Figure 1, Table 1**). Thus, our model is robustly predictive of the expanding separation in mortality per capita between BCG and non-BCG vaccinating countries over time.

### Confounding factors

A number of additional outliers and confounding factors exist, known and unknown (See Supplemental Notes for a detailed analysis of multiple confounding factors). Whilst we ensure that our analysis is subject to rigorous empiricism using noise-free higher quality data, it is impossible to know all the confounding possibilities, for which clinical trials and more high-quality temporal case and mortality data should provide more insight in the future. Important confounding factors in our study included the variabilities in detection of cases, the temporal nature of the virus spread, the influence of dynamic populations, age stratification demographics, urbanization, wealth of a country, rates of testing, variable death certification and reporting times as well as some differences between the planned and actual BCG execution within countries. However, many of these confounding factors were not exhaustively analyzed and should be explored in greater detail, and in particular *quantified*, in further work. Importantly, we have sought to address each one of the aforementioned confounders in the Supplemental Notes accompanying this paper. Each has been carefully analyzed and two of the major confounders are examined in the following sections. Finally, strain type, which is often dynamic over time was collected in some cases but has not yet been modelled, due to non systematic collection of this data in most of the world.

### Confounding factors: high testing rates

Recent studies^7^ have asserted that variable testing regimes reveal distorted correlations between BCG vaccination and COVID19 mortality rates. Where countries performing numbers of tests for COVID19 result in high numbers of cases and mortality. These studies use three types of analyses: cases, cases per million, and percentage mortality per total cases. These approaches at their core depend on case data which does indeed skew mortality in a similar way as case fatality rate (CFR). Therefore all these analyses inherently suffer from the absence of a comparable variable across countries. Nevertheless, we tested this assertion against our model. If the assertion is correct the top 20 high testing countries (as reported) would be composed almost entirely of non BCG vaccinating nations. Instead both BCG and non BCG vaccinating countries were well represented among the top 20 “testing countries”, with mortality rates diverging as before. See (**Figure 2)**. This analysis suggests that testing “bias” from countries performing high numbers of tests does not skew mortality rates of BCG versus nonBCG vaccinating countries.

**Figure 2.**
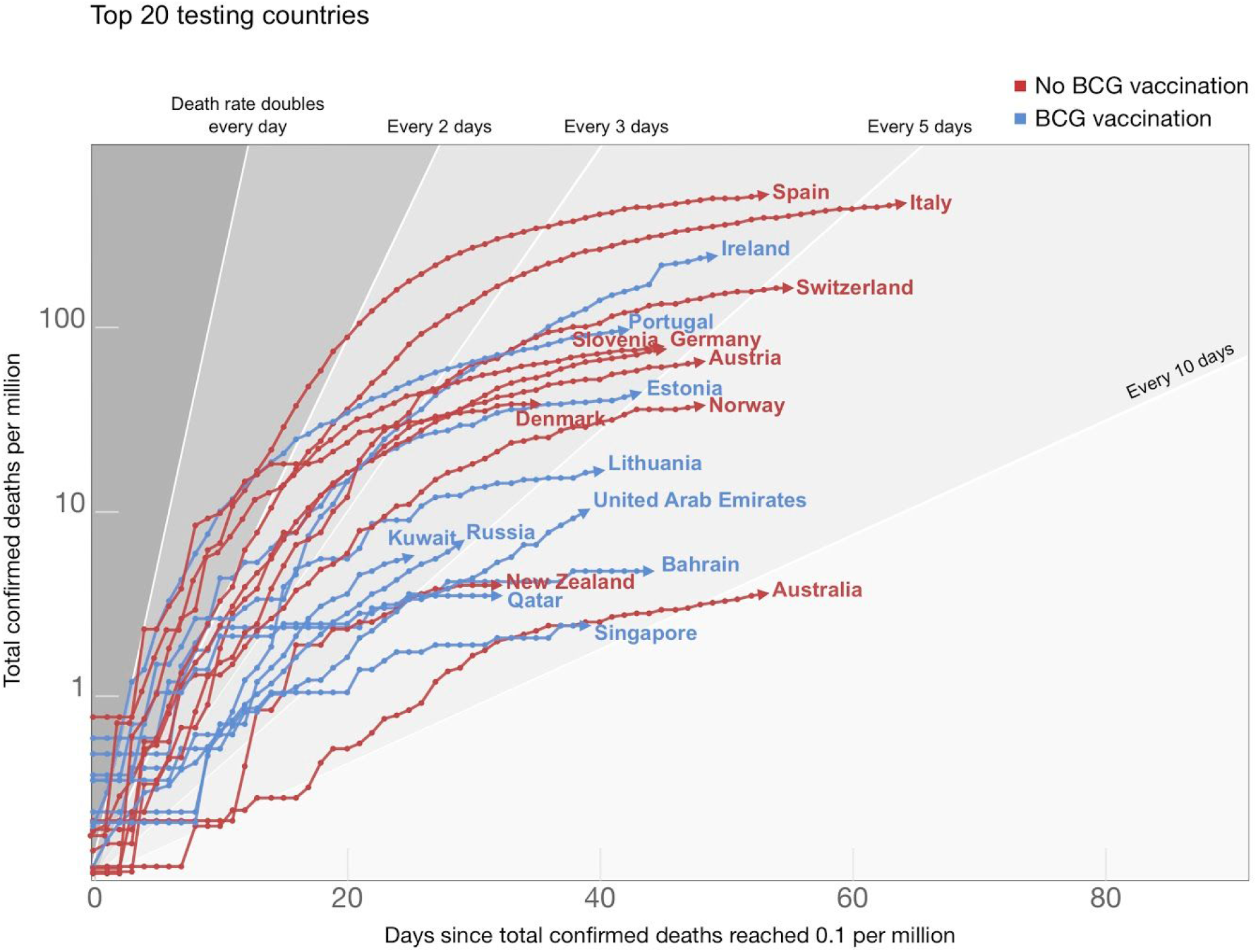
Both BCG and non BCG vaccinating countries are well represented among the top 20 “testing countries”. No separation can be drawn due to the frequency of testing by either group. Data represents days since the total confirmed deaths of COVID-19 per million people reached 0.1. The variable time span is from 31 December 2019 to 31 April 2020. Source data^4^.

### Confounding factors: age as a confounder

Age as a confounder. A leitmotif of COVID-19 is the much higher risk of fatality that occurs in older aged individuals. A study from Japan controlling for life expectancy finds that “this BCG effect remained significant after controlling for the country’s life expectancy”.^8^ Conversely, fatalities around the world are rare in children below the age of 10 yrs old. Children have been identified as major spreaders of influenza.^9^ In the current COVID-19 pandemic our analysis of BCG versus nonBCG vaccinating countries cannot reveal the role of children since there is insufficient data to examine fatalities in the age group. However, there are hints in the data that populations where young people are routinely vaccinated whilst older populations have been unvaccinated have a slower growth in fatalities. Out of all the countries that have ever routinely administered BCG to children there are only a few who have given up vaccinating completely from one year to the next in the last 20 or so years. We can use this limited data to look at what happens when only children are excluded from BCG in a country, while those who are older still have coverage. In Finland, Austria, Norway, Czech Republic and Slovenia children no longer get routine BCG but those who are today in their early teens are well covered. Contrasting these countries with Ireland and the UK where only those in their late 20’s and up are actually covered shows an interesting separation and comparing both groups to the control group Japan, Qatar, Singapore, Thailand where all children are covered also furthers this interesting study (**Figure 3**). With more data over time these effects will be more easily quantified.

**Figure 3.**
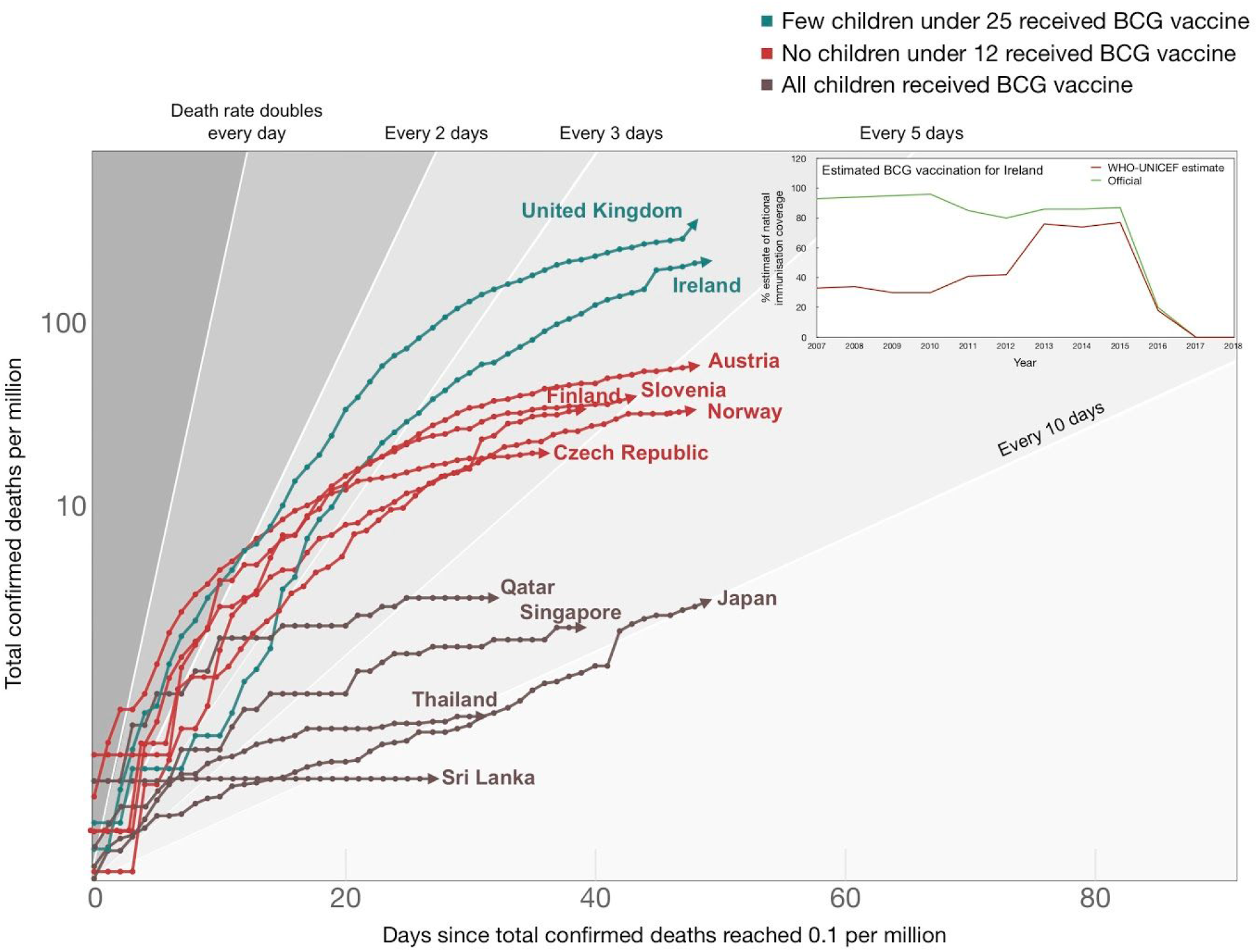
Countries that routinely vaccinate all children exhibit reduced COVID-19-associated mortality rates. Although every child in Ireland is reported to receive the BCG vaccine, there are large disparities between Official (Irish government) reported BCG vaccination compliance and estimated vaccine coverage reported by the WHO using actual vaccine batches (INSET). Data represents days since the total confirmed deaths of COVID-19 per million people reached 0.1. The variable time span is from 31 December 2019 to 31 April 2020. Source data^4^.

### Correlation not causation: *whether* vs *how much* ?

Importantly, this analysis at this early stage can show predictive correlations, but alone cannot assign causation. These variables and the sheer paucity of data preclude our analytical approach from indicating “*how much*” of an influence BCG could play in a quantitative manner at this time, though possibly with more data that could soon change. In contrast, our early analysis could indicate “*whether*” BCG has an influence. In time and with more data points, stronger empirical evidence will emerge that can bring more quantitative information. Notwithstanding, clear strong signals have emerged especially in the dynamic mortality per capita data as to “*whether*” BCG is influential and given our current situation, this is worth discussing expediently.

### Observations of non-specific effects of BCG

The BCG vaccine is primarily used against tuberculosis (TB).^10^ *Mycobacterium tuberculosis* (Mtb), the bacteria that causes TB, currently infects approximately one quarter of the world’s population. TB is associated with high morbidity and mortality, particularly in Sub-Saharan Africa. In 2018, 10 million cases of tuberculosis (TB) and over 1.5 million TB-associated deaths were recorded^11^, which makes it the deadliest infectious disease on the planet. Numerous studies have investigated whether BCG vaccination reduces TB disease burden and associated mortality. Neonatal BCG vaccination has been shown to be associated with lower mortality rates among both TB-exposed and TB-unexposed children. ^12^ Three randomized controlled trials of early BCG vaccination in Guinea-Bissau found a 38% reduction in all-cause neonatal mortality. ^13^ These effects do not only benefit infants, as a recent clinical trial revealed that revaccinating at-risk teenagers with the BCG vaccine they first got as infants, could significantly reduce the threat of these patients becoming carriers of TB infection. ^1^ This latter study has also demonstrated a 73% decrease in non-mycobacterial respiratory tract infections induced by BCG vaccination (data collected within the safety monitoring plan, as the main endpoint of the study was tuberculosis infection). Collectively, these studies provide evidence that the BCG vaccine provides broad immune protective properties against non-related infections and disease. Several studies strongly support this hypothesis. For example, a randomized placebo-controlled study showed that BCG vaccination lowers yellow fever vaccine viremia. ^14^ In addition, BCG has been shown to offer protection against bladder cancer ^15^ and malaria. ^16^ Overall, these studies indicate that the strong protective role of the BCG vaccine is not only limited to TB and indicate that interventions to increase timely BCG vaccination are urgently warranted.

### What is the mechanism by which BCG can confer this non-specific immunity?

The heterologous effects of BCG vaccination also served as the basis for the discovery of ‘trained immunity’, the *de facto* immune memory of the innate immune system. ^17^ Innate immune cells (such as monocytes) exposed to BCG or other stimuli (e.g. β-glucan) prior to exposure to certain infectious agents (e.g. TB), developed enhanced resistance upon reinfection with a second infectious agent, a response referred to as ‘trained immunity’.^17, 18^ The enhanced responsiveness observed in trained monocytes is accompanied by durable metabolic changes and the stable accumulation of epigenetic marks on dozens of innate immune responsive genes and enhancer elements. 18-21 As a consequence of epigenetic reprogramming, trained genes are more strongly and robustly transcribed. Critically, quantitative correlates of trained immunity beyond increases in deposition of epigenetic marks that can be rapidly and easily interrogated, are lacking.

Epigenetic modifications are catalyzed by a large family of chromatin remodelers. ^22^ However, the precise mechanism describing how epigenetic marks are directed to specific regions of the genome has been lacking. Accumulating evidence reveals the ‘non coding’ portion of the genome has a significant role in gene regulation. ^23,24^ Recently, it has been shown that the non coding genome is extensively transcribed into long non-coding RNA (lncRNA).^25, 26^ LncRNAs can use the folding of DNA in 3D to bring chromatin remodelers proximal to target genes to regulate the epigenetic activation of genes. ^27^ A new class of lncRNAs, called Immune Priming LncRNAs (IPLs) have been shown to regulate the “writing” of innate immune memory at “trained” immune genes.^28^ Importantly, assaying the levels of epigenetic modifications at innate immune genes may represent a novel approach to assess the degree of “trained” immune activation in BCG vaccinated individuals. The ability to routinely assay such levels in patients post-vaccination (Moorlag et al, in revision) would have significant benefits in assessing vaccine efficacy as well as nonspecific benefits, such as the reduction of respiratory infections unrelated to Mtb.

Until recently, it has been unclear how this vaccine is able to generate long-term innate immune memory. Recently, Kaufmann et al.,^29^ used a mouse model to show that BCG reprograms hematopoietic stem cells (HSC) and multipotent progenitors (MPPs) in the bone marrow (**Figure 4**). Notably, this leads to the expansion and polarisation of MPPs towards myelopoiesis. Further, the macrophages that arise from BCG-reprogrammed HSCs are epigenetically reprogrammed and greatly enhanced in their ability to offer protection to Mtb infection.^29^ This infers that the BCG vaccination-induced epigenetic changes in HSCs and/or MPPs can be transferred to circulating monocytes, and subsequently macrophages. This process enables myeloid cells to preserve a memory of exposure to pathogens, leading to long-lasting enhanced protection against infectious agents, such as Mtb. Similar processes have been described in the bone marrow of human volunteers vaccinated with BCG, in which transcriptional and epigenetic reprogramming of HSCs has been documented three months after the vaccination (Cirovic et al, in revision). Interestingly, in humans it has been demonstrated that not only monocytes/macrophages, but also the neutrophils display a long-term functional boost after BCG vaccination (Moorlag et al, in revision).

**Figure 4:**
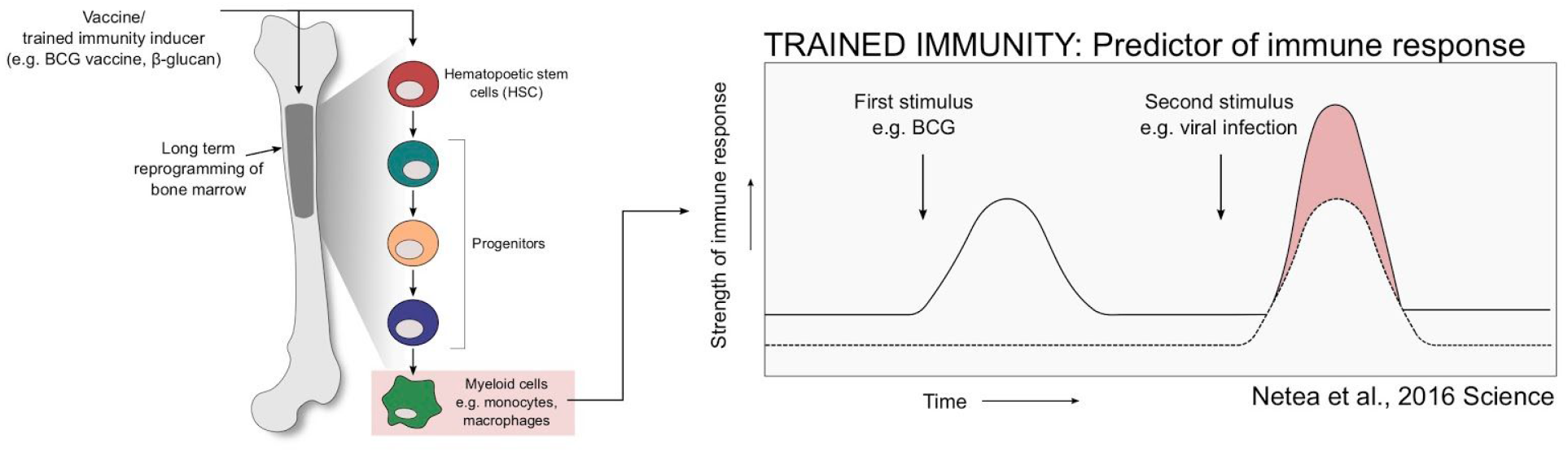
Stimuli that induce trained immunity (e.g. BCG) reprogram the bone marrow niche to induce long term immune memory. As a consequence, myeloid cells that arise from the bone marrow are enhanced in their capability to clear infection.

## DISCUSSION

With the rapid spread of SARS-CoV-2 around the world, an outstanding question is whether BCG vaccination will combat the symptoms and reduce the overall mortality rate of the respiratory disease caused by this virus. The ongoing COVID-19 pandemic represents a global ‘uncontrolled clinical trial’ of BCG vaccinated versus unvaccinated populations coupled to vaccine strain. However, validating the suggested effect of BCG vaccination on COVID-19 in randomized clinical trials is a crucial step before being able to conclude that this approach should be expanded as a tool in this pandemic. It was important to perform our study early in the pandemic, not only to support trials or explore an early prophylactic, but also as an important opportunity to understand correlations with BCG before suppression techniques influenced the data. This could mean, for example, that while a large amount of data emerges over time, a very important set of data quality snapshots in time could only be acquired at the very early stages of the pandemic for each country as these would be less influenced by the divergent suppression actions each jurisdiction has taken. Notwithstanding the above, our sample set of countries early on in the pandemic did contain many examples of high and low suppression countries with high and low fatalities and both high and low BCG vaccination rates that strongly correlate with BCG vaccination protecting from mortality in populations infected with COVID-19. This model can be replicated at multiple “time stamps” (**Table 1**) by any party with access to identical publicly available data.

Many studies of the type we have undertaken are termed “ecological”, with the incumbent prejudices that come with the label. Indeed ecological studies often suffer from confounding factors and outlying data that most models stretch to accommodate. Generally two claims are made to weaken ecological studies. Firstly, that conclusions are inferred about individuals from the results of aggregate data, and secondly, that there is an inability to adequately control for confounders. We avoided these two pitfalls by using highly robust and cross-checked *country specific* actual immunization history along with actual *country specific* deaths from COVID-19. Though we do make use of individual and aggregate data, we have never combined the two datasets. Where possible we have taken a data quality approach to estimate *individual make up of the aggregate*. An example, using detailed age stratifications and migration numbers in analyzing outliers such as in Lebanon (see Supplemental Notes). We go to great lengths to make sure confounders are systematically and rigorously covered by way of Supplemental Notes and further data analysis conducted on each specific confounding variable to be verified against our model. There is an ever increasing quantity of pandemic data available, of highly variable quality. We cannot emphasize enough that the use of data-quality driven data science to tease out hints from patterns has been a cornerstone of our approach.

The exact duration of protection from TB and other diseases by BCG remains unknown. A study conducted in American Indians and Alaska Natives revealed that BCG vaccine efficacy against tuberculosis persisted for 50 - 60 years. ^30^ Interestingly, this study revealed a moderate reduction in the efficacy of the vaccine over time, the effect of which was more pronounced in men. A retrospective cohort study in Norwegian-born individuals vaccinated with BCG showed an average vaccine effectiveness against all forms of tuberculosis of 49% in a 40-year follow-up. ^31^ Overall, this indicates that a single dose of BCG vaccine can have a long duration of protection against TB. Yet, the induction of non-specific protection against heterologous infections is however likely to be shorter-lived, although this remains to be demonstrated. As a consequence, it is unlikely that children receiving the BCG vaccine many decades ago will be protected from COVID-19 now. For example, this may be due to the administration of additional live vaccines, a subject of intense investigation. More relevant to this study, children have been identified as the major spreaders of influenza, therefore it is tempting to speculate that they may also be partly responsible for enhancing the SARS-CoV-2 community spread of SARS-CoV-2. Recent data indicates children can be asymptomatic yet harbor amounts of virus equivalent or exceeding adults. ^32^ Hints of this may be apparent in our analysis (**Figure 3**). As a consequence of enhanced innate immune responses, children vaccinated with BCG may be more likely to rapidly clear SARS-CoV-2. In turn, this may reduce the community spread of SARS-CoV-2, which reduces the overall disease burden especially in more vulnerable segments of the population.

Currently, there are several prospective trials being conducted or being prepared worldwide (including ongoing studies in The Netherlands, Australia and Greece) to investigate whether the BCG vaccine would be effective against COVID-19. We will only know the outcome of these trials in several months and the BCG vaccine is in short supply. Therefore, in order to ensure that children in high-risk areas receive protection against TB, it is important that the BCG vaccine is only used in these controlled trials for the moment. In addition, it is vital to carefully assess safety data from these clinical trials to ensure that the enhanced innate immunity induced by BCG does not exacerbate COVID-19, especially in patients with underlying immune conditions.

Clearly, the role of trained immunity in mediating the protective effects of the BCG vaccine warrants further investigation. Several avenues of enhancing trained immune responses have been identified (reviewed in Mulder et al., 2019).^33^ These novel approaches could well serve as a simple and effective measure in reducing COVID-19 associated mortality. Critically, these approaches can be implemented prior to the generation of a SARS-CoV2 vaccine, a development that at its very earliest is over a year away and its long term efficacy remains uncertain.

## Data Availability

Data available upon request. All COVID-19 data is already in the public domain.

https://ourworldindata.org/coronavirus

## Acknowledgments

We thank all members of the Joosten, Netea and Mhlanga laboratories for their comments on this manuscript. We also thank all the clinicians, physicians and medical workers around the world who have collected the vast majority of this data.

## References

1. Nemes E, Geldenhuys H, Rozot V, Rutkowski KT, Ratangee F, Bilek N, et al. Prevention of M. tuberculosis Infection with H4:IC31 Vaccine or BCG Revaccination. N Engl J Med. 2018;379(2):138–49.

2. Moorlag S, Arts RJW, van Crevel R, Netea MG. Non-specific effects of BCG vaccine on viral infections. Clin Microbiol Infect. 2019;25(12):1473–8.

3. Qasim B, Yusuf J. Will Coronavirus Pandemic Diminish by Summer? Available at SSRN: https://ssrncom/abstract=3556998. 2020.

4. Roser M, Ritchie H, Ortiz-Ospina E. Coronavirus Disease (COVID-19) – Statistics and Research. Published online at OurWorldInDataorg Retrieved from: ‘https://ourworldindataorg/coronavirus’ [Online Resource]. 2020.

5. COVID-19 Coronavirus Pandemic. Worldometers.

6. Leung K, Wu JT, Liu D, Leung GM. First-wave COVID-19 transmissibility and severity in China outside Hubei after control measures, and second-wave scenario planning: a modelling impact assessment. Lancet. 2020;395(10233):1382–93.

7. Hensel J, McGrail DJ, McAndrews KM, Dowlatshahi D, LeBleu VS, Kalluri R. Exercising caution in correlating COVID-19 incidence and mortality rates with BCG vaccination policies due to variable rates of SARS CoV-2 testing. *medRxiv*. 2020:2020.04.08.20056051.

8. Sala G, Miyakawa T. Association of BCG vaccination policy with prevalence and mortality of COVID-19. *medRxiv*. 2020:2020.03.30.20048165.

9. Heikkinen T. Influenza in children. Acta Paediatrica. 2006;95(7):778–84.

10. Zwerling A, Behr MA, Verma A, Brewer TF, Menzies D, Pai M. The BCG World Atlas: a database of global BCG vaccination policies and practices. PLoS Med. 2011;8(3):e1001012.

11. Global tuberculosis report. Geneva: World Health Organization. 2019.

12. Thysen SM, Benn CS, Gomes VF, Rudolf F, Wejse C, Roth A, et al. Neonatal BCG vaccination and child survival in TB-exposed and TB-unexposed children: a prospective cohort study. BMJ Open. 2020;10(2):e035595.

13. Jensen KJ, Biering-Sorensen S, Ursing J, Kofoed PL, Aaby P, Benn CS. Seasonal variation in the non-specific effects of BCG vaccination on neonatal mortality: three randomised controlled trials in Guinea-Bissau. BMJ Glob Health. 2020;5(3):e001873.

14. Arts RJW, Moorlag SJCFM, Novakovic B, Li Y, Wang S-Y, Oosting M, et al. BCG Vaccination Protects against Experimental Viral Infection in Humans through the Induction of Cytokines Associated with Trained Immunity. Cell Host & Microbe. 2018;23(1):89–100.e5.

15. Buffen K, Oosting M, Quintin J, Ng A, Kleinnijenhuis J, Kumar V, et al. Autophagy controls BCG-induced trained immunity and the response to intravesical BCG therapy for bladder cancer. PLoS Pathog. 2014;10(10):e1004485.

16. Berendsen ML, van Gijzel SW, Smits J, de Mast Q, Aaby P, Benn CS, et al. BCG vaccination is associated with reduced malaria prevalence in children under the age of 5 years in sub-Saharan Africa. BMJ Glob Health. 2019;4(6):e001862.

17. Kleinnijenhuis J, Quintin J, Preijers F, Joosten LA, Ifrim DC, Saeed S, et al. Bacille Calmette-Guerin induces NOD2-dependent nonspecific protection from reinfection via epigenetic reprogramming of monocytes. Proc Natl Acad Sci U S A. 2012;109(43):17537–42.

18. Quintin J, Saeed S, Martens JHA, Giamarellos-Bourboulis EJ, Ifrim DC, Logie C, et al. Candida albicans infection affords protection against reinfection via functional reprogramming of monocytes. Cell Host Microbe. 2012;12(2):223–32.

19. Cheng SC, Quintin J, Cramer RA, Shepardson KM, Saeed S, Kumar V, et al. mTOR- and HIF-1alpha-mediated aerobic glycolysis as metabolic basis for trained immunity. Science. 2014;345(6204):1250684.

20. Saeed S, Quintin J, Kerstens HH, Rao NA, Aghajanirefah A, Matarese F, et al. Epigenetic programming of monocyte-to-macrophage differentiation and trained innate immunity. Science. 2014;345(6204):1251086.

21. Arts RJ, Novakovic B, Ter Horst R, Carvalho A, Bekkering S, Lachmandas E, et al. Glutaminolysis and Fumarate Accumulation Integrate Immunometabolic and Epigenetic Programs in Trained Immunity. Cell Metab. 2016;24(6):807–19.

22. Langst G, Manelyte L. Chromatin Remodelers: From Function to Dysfunction. Genes (Basel). 2015;6(2):299–324.

23. Amaral PP, Dinger ME, Mercer TR, Mattick JS. The eukaryotic genome as an RNA machine. Science. 2008;319(5871):1787–9.

24. Sanjana NE, Wright J, Zheng K, Shalem O, Fontanillas P, Joung J, et al. High-resolution interrogation of functional elements in the noncoding genome. Science. 2016;353(6307):1545–9.

25. Iyer MK, Niknafs YS, Malik R, Singhal U, Sahu A, Hosono Y, et al. The landscape of long noncoding RNAs in the human transcriptome. Nat Genet. 2015;47(3):199–208.

26. Hon CC, Ramilowski JA, Harshbarger J, Bertin N, Rackham OJ, Gough J, et al. An atlas of human long non-coding RNAs with accurate 5’ ends. Nature. 2017;543(7644):199–204.

27. Wang KC, Yang YW, Liu B, Sanyal A, Corces-Zimmerman R, Chen Y, et al. A long noncoding RNA maintains active chromatin to coordinate homeotic gene expression. Nature. 2011;472(7341):120–4.

28. Fanucchi S, Fok ET, Dalla E, Shibayama Y, Borner K, Chang EY, et al. Immune genes are primed for robust transcription by proximal long noncoding RNAs located in nuclear compartments. Nat Genet. 2019;51(1):138–50.

29. Kaufmann E, Sanz J, Dunn JL, Khan N, Mendonca LE, Pacis A, et al. BCG Educates Hematopoietic Stem Cells to Generate Protective Innate Immunity against Tuberculosis. Cell. 2018;172(1-2):176-90 e19.

30. Aronson NE, Santosham M, Comstock GW, Howard RS, Moulton LH, Rhoades ER, et al. Long-term efficacy of BCG vaccine in American Indians and Alaska Natives: A 60-year follow-up study. JAMA. 2004;291(17):2086–91.

31. Nguipdop-Djomo P, Heldal E, Rodrigues LC, Abubakar I, Mangtani P. Duration of BCG protection against tuberculosis and change in effectiveness with time since vaccination in Norway: a retrospective population-based cohort study. Lancet Infect Dis. 2016;16(2):219–26.

32. Jones TC, Mühlemann B, Veith T, Zuchowski M, Hofmann Jr, Edelmann AS, et al. An analysis of SARS-CoV-2 viral load by patient age. https://zoonosencharitede/fileadmin/user_upload/microsites/m_cc05/virologie-ccm/dateien_upload/Weitere_Dateien/analysis-of-SARS-CoV-2-viral-load-by-patient-agepdf.

33. Mulder WJM, Ochando J, Joosten LAB, Fayad ZA, Netea MG. Therapeutic targeting of trained immunity. Nat Rev Drug Discov. 2019;18(7):553–66.

